# Age Related Differences in BMD Response During Three Years of Denosumab Treatment

**DOI:** 10.64898/2026.05.25.26354051

**Authors:** Koji Ishikawa, Tomoyuki Asada, William Richardson, Choiselle Marius, Mahoko Ishikawa, Tuyet Nguyen, Philip Varnadore, Soji Tani, Peter Passias, Benjamin Aaron Alman

## Abstract

**Introduction:** Denosumab increases bone mineral density and reduces fracture risk in patients with osteoporosis. However, whether BMD response to denosumab differs by age, particularly during longer term treatment, remains unclear. This study investigated the association between baseline age and BMD gain during 3 years of denosumab treatment in patients with osteoporosis.

**Methods:** This retrospective study included patients with osteoporosis who were treated with denosumab. DXA-based BMD and bone turnover markers were followed for up to 3 years. Percent BMD gain from baseline, defined as %BMD gain, was evaluated. The longitudinal association between baseline age and %BMD gain was assessed using multivariable linear mixed-effects models for the lumbar spine and total hip. Analyses were performed in the treatment naive cohort and the overall cohort according to prior osteoporosis treatment status.

**Results:** A total of 255 patients were included in the analysis, of whom 110 had not received prior osteoporosis treatment. In multivariable linear mixed-effects models, older baseline age was associated with smaller lumbar spine %BMD gain in the treatment naive cohort at both 1 and 3 years. Each 1-year increase in age was associated with a 0.187 percentage-point lower lumbar spine %BMD gain at 1 year and a 0.293 percentage-point lower gain at 3 years (1 year: β = −0.187, p = 0.006, 3 years: β = −0.293, p = 0.031). In contrast, baseline age was not significantly associated with total hip %BMD gain in the treatment naive cohort (1 year: β = −0.011, p = 0.826; 3 years: β = 0.028, p = 0.727). In the overall cohort, baseline age was not significantly associated with %BMD gain at either the lumbar spine or total hip at 1 or 3 years (all p > 0.05).

**Conclusion:** Older baseline age was associated with a modestly smaller lumbar spine BMD gain in treatment naive patients, whereas no significant age-related association was observed at the total hip. In the overall cohort, age was not significantly associated with BMD gain at either site. These findings suggest that age may have a limited, site specific influence on BMD response to denosumab, particularly in treatment naive patients, and may support more individualized treatment planning in patients with osteoporosis.

## Introduction

Denosumab is a monoclonal antibody against receptor activator of nuclear factor κB ligand that suppresses osteoclast mediated bone resorption. In postmenopausal women with osteoporosis, denosumab increases bone mineral density and reduces the risk of fractures^1,2^. Long term extension studies have further shown progressive BMD gains at both the lumbar spine and hip for up to 10 years^3,4^. However, the effects of denosumab are reversible after discontinuation, and treatment withdrawal can lead to excessive bone resorption, loss of acquired BMD, and an increased risk of multiple vertebral fractures^5–7^. Thus, denosumab initiation requires careful consideration of patient age, as age may influence the expected duration of treatment and the need for subsequent transition therapy.

Aging is accompanied by multiple skeletal changes, including reduced bone formation capacity, increased cortical porosity, and different patterns of trabecular and cortical bone loss across skeletal sites^8–10^. These skeletal alterations are associated with changes in bone remodeling, with a shift toward osteoclast mediated resorption relative to bone formation^11^. As a result, the skeletal response to osteoporosis treatment may vary according to patient age and anatomical site. Although antiosteoporosis medications generally remain effective in older adults, previous studies have reported inconsistent findings regarding age related differences in BMD response and fracture outcomes^12–16^.

This question may be particularly relevant for denosumab because its therapeutic effects are mediated through direct inhibition of RANKL dependent osteoclast activity^1,4^. By suppressing osteoclast mediated bone resorption, denosumab produces sustained increases in BMD, but the magnitude of these gains may depend on the remodeling environment at treatment initiation. Therefore, age related differences in skeletal remodeling activity may influence the longitudinal BMD response to denosumab. Prior subgroup analyses of the FREEDOM trial have shown that denosumab reduces fracture risk in older women, including those aged 75 years or older, and improves BMD in this age group^17,18^. However, the association between baseline age and BMD gains during longer term denosumab treatment in routine clinical practice remains unclear.

Clarifying this relationship may provide clinically useful information when considering denosumab initiation and long-term treatment strategies across different age groups. In this retrospective study, we investigated whether baseline age was associated with BMD response during 3 years of denosumab treatment in treatment naive patients with osteoporosis.

## Materials and Methods

### Study design and patient selection

This retrospective observational study included patients with primary osteoporosis in Japan who were treated with denosumab. The study was approved by the institutional ethics committee (No. 17091229-5) and was conducted in accordance with the principles of the Declaration of Helsinki. The requirement for written informed consent was waived because of the retrospective nature of the study.

Patients were eligible if they had baseline DXA data and at least one follow up DXA measurement within 3 years after denosumab initiation. Patients were excluded if baseline DXA data were unavailable, if no follow up DXA data were available during the 3 year follow up period, or if they had secondary osteoporosis, poorly controlled thyroid disease, active malignancy, or conditions that could substantially affect mobility or skeletal health, such as Parkinson’s disease. Because prior osteoporosis treatment may influence subsequent BMD response to denosumab, patients with no prior osteoporosis medication before denosumab initiation were defined as the treatment naive cohort and used for the primary analysis^19–21^. The overall cohort included patients regardless of prior osteoporosis treatment status and was analyzed as a secondary cohort.

### Data collection

Baseline demographic and clinical variables included age, sex, body mass index (BMI, kg/m²), smoking and alcohol. Laboratory variables assessed before treatment included serum calcium (mg/dL), phosphorus (mg/dL), estimated glomerular filtration rate (eGFR, mL/min/1.73 m²) and alkaline phosphatase (ALP, U/L). Serum bone turnover markers (BTMs) included tartrate-resistant acid phosphatase 5b (TRACP-5b; reference range in women, 120–420 mU/dL; measured using the Osteolinks® TRACP-5b® test kit, DS Pharma Biomedical Co., Ltd., Osaka, Japan) and total N-terminal propeptide of type I procollagen (total P1NP; reference range in postmenopausal women, 26.4–98.2 μg/L; measured using a total P1NP assay on an Elecsys automated analyzer, Roche Diagnostics, Switzerland). BTMs were assessed at baseline and repeatedly during follow-up, including 6 months, 12 months, 24 months, and 36 months after denosumab initiation.

Dual energy X ray absorptiometry (DXA) of the lumbar spine and total hip was performed at baseline and annually for up to 3 years after denosumab initiation. Areal BMD of the lumbar spine (L1–L4) and total hip was measured using DXA (Hologic QDR series; Hologic, Waltham, MA, USA). All DXA measurements were analyzed centrally by a radiologist. The primary longitudinal outcome was percent BMD gain from baseline (%BMD gain), calculated as 100 × (follow-up BMD − baseline BMD) / baseline BMD.

### Statistical analysis

All statistical analyses were performed using R software (version 4.4.3; R Core Team, Vienna, Austria). Continuous variables were summarized as means with standard deviations. Categorical variables were summarized as counts and percentages. A two-sided p value <0.05 was considered statistically significant. Univariate comparisons across age groups were performed using one-way ANOVA for continuous variables and chi-square tests for categorical variables.

Multivariable linear mixed-effects models were used to examine the association between baseline age as a continuous variable and %BMD gain at 1 and 3 years after denosumab initiation. Follow-up time was modeled using natural cubic splines, and an interaction term between baseline age and follow-up time was included to estimate time-specific associations between age and %BMD gain. Models were constructed separately for lumbar spine BMD and total hip BMD. Covariates included sex, BMI, eGFR, and prior osteoporosis treatment status in the overall cohort. The overall age-by-time interaction was evaluated using Type III analysis of variance with Satterthwaite’s approximation for degrees of freedom. In the treatment naive cohort, prior osteoporosis treatment status was not included as a covariate. Models included patient-specific random intercepts and random slopes for follow-up time. Model-based estimates were used to calculate the adjusted difference in %BMD gain per 1-year increase in baseline age at 1 and 3 years.

To complement the above analysis, patients were categorized into age groups based on cohort specific quartiles: ≤72, 72 to 77, 77 to 83, and ≥83 years. Age group specific trajectories of %BMD gain through 3 years were modeled using natural spline based linear mixed effects models, providing model-based visualization of longitudinal BMD response patterns across age groups. Bone turnover markers were also summarized descriptively by age group at baseline, 6 months, and 1, 2, and 3 years after denosumab initiation, without formal hypothesis testing.

## Results

### Baseline patient characteristics

Between January 2015 and January 2023, 761 patients who initiated osteoporosis treatment were screened. Of these, 338 patients treated with denosumab were assessed for eligibility, and 83 were excluded, including 48 with unavailable baseline data and 35 with no follow up DXA data during the 3 year follow up period. The remaining 255 patients were included in the overall cohort for secondary analysis. Among them, 110 treatment naive patients were included in the primary analysis (Fig. 1).

**Figure 1.**
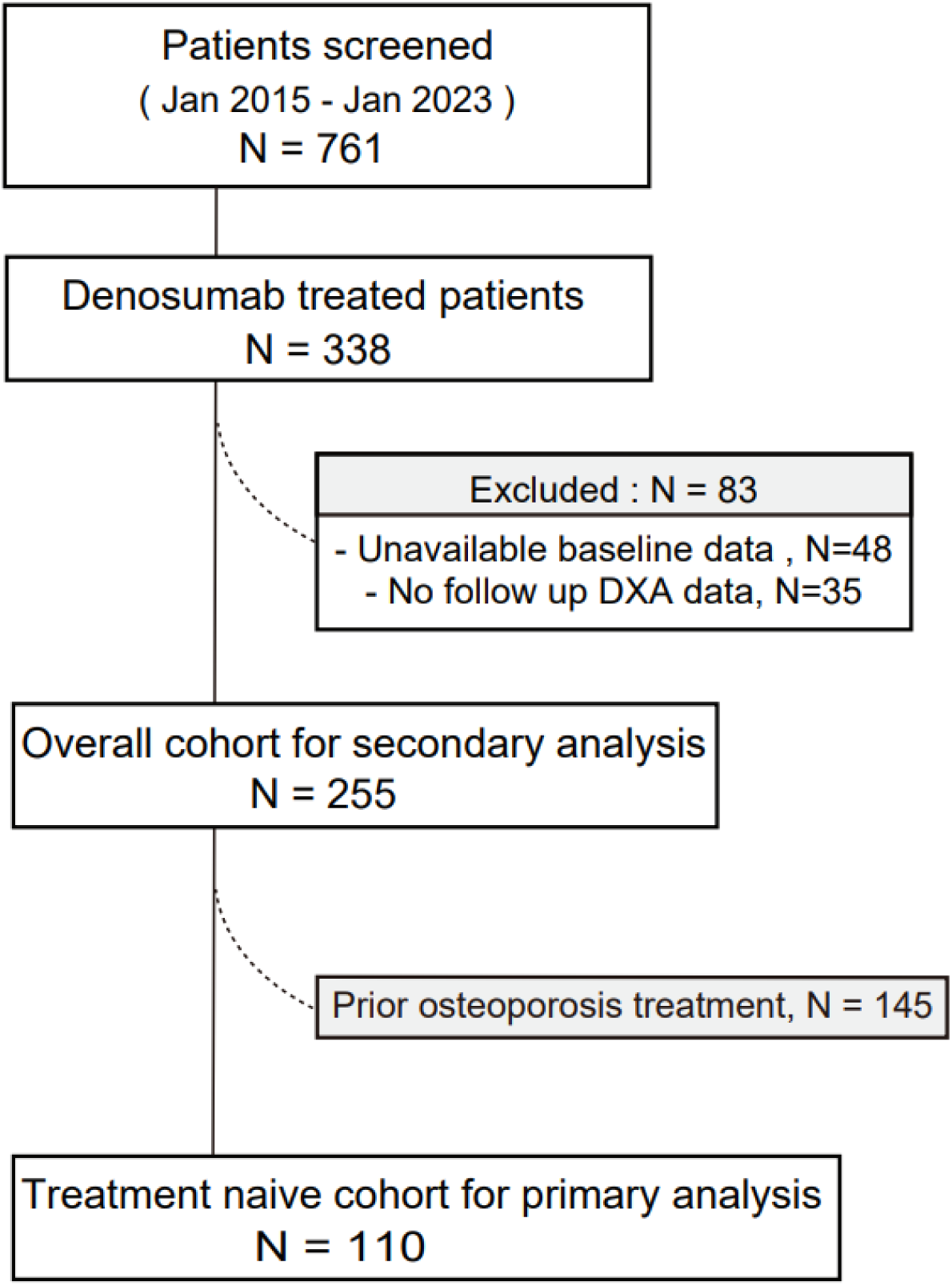
Patient flow diagram. Patients who initiated osteoporosis treatment between January 2015 and January 2023 were screened. Among 761 screened patients, 338 patients treated with denosumab were assessed for eligibility. Eighty-three patients were excluded because of unavailable baseline data or no follow up DXA data. The remaining 255 patients were included in the overall cohort for secondary analysis, and 110 treatment naive patients were included in the primary analysis.

In the treatment naïve patients, there were no significant differences in baseline characteristics across age groups, except for total hip BMD (Table 1). In the overall cohort, although prior osteoporosis treatment did not differ significantly across age groups (p = 0.30), eGFR was lower in the older age groups (p < 0.001). Spine BMD was similar among age groups, whereas total hip BMD differed significantly across age groups and was lowest in the oldest group (≥83 years, 0.558 g/cm²; p < 0.001) (Supplementary Table 1). All but five patients received supplemental vitamin D to prevent hypocalcemia^22^.

**Table 1.** Demographic Data in treatment naïve cohort. TRACP-5b = Tartrate-resistant acid phosphatase-5b; P1NP = procollagen type 1 N-terminal propeptide; IP = inorganic phosphate; ALP = Alkaline phosphatase; Adjusted Ca=albumin-corrected calcium; DXA = Dual-energy X-ray absorptiometry; BMD = bone mineral density

|  | **Overall** | **≤ 72** | **72-77** | **77-83** | **≥ 83** | **p** |
| --- | --- | --- | --- | --- | --- | --- |
| **n** | 110 | 36 | 24 | 27 | 23 |  |
| **Age, year (mean (SD))** | 76.3 (8.3) | 67.4 (5.3) | 74.5 (1.5) | 80.1 (1.8) | 87.6 (3.0) | <0.001 |
| **Male, n(%)** | 10 (9.1) | 3 (8.3) | 4 (16.7) | 2 (7.4) | 1 (4.3) | 0.492 |
| **BMI, kg/m<sup>2</sup> (mean (SD))** | 22.3 (4.7) | 23.6 (4.8) | 21.4 (6.4) | 21.8 (3.6) | 22.0 (3.6) | 0.438 |
| **Smoking, n(%)** | 2 (1.8) | 1 (2.9) | 1 (4.2) | 0 (0.0) | 0 (0.0) | 0.642 |
| **Alcohol, n(%)** | 2 (1.8) | 0 (0.0) | 1 (4.2) | 1 (3.7) | 0 (0.0) | 0.503 |
| **Blood test** |  |  |  |  |  |  |
| **eGFR, mL/min/1.73m<sup>2</sup> (mean (SD))** | 65.1 (19.4) | 71.4 (18.3) | 65.5 (12.6) | 61.0 (16.7) | 59.6 (26.9) | 0.073 |
| **Adjusted Ca, mg/dL (mean (SD))** | 9.3 (0.4) | 9.4 (0.3) | 9.2 (0.3) | 9.2 (0.5) | 9.3 (0.5) | 0.324 |
| **IP, mg/dL (mean (SD))** | 3.6 (0.4) | 3.7 (0.5) | 3.6 (0.4) | 3.5 (0.3) | 3.5 (0.5) | 0.469 |
| **ALP, U/L (mean (SD))** | 297.8 (105.3) | 306.0 (100.5) | 292.5 (83.5) | 301.4 (125.1) | 286.2 (112.9) | 0.901 |
| **TRACP-5b, mU/dL (mean (SD))** | 537.0 (197.1) | 500.1 (165.8) | 556.5 (191.2) | 557.0 (238.8) | 552.0 (198.9) | 0.599 |
| **Total P1NP, µg/L (mean (SD))** | 83.5 (66.1) | 71.0 (32.6) | 72.9 (41.4) | 98.1 (78.8) | 96.5 (99.6) | 0.254 |
| **DXA measurement** |  |  |  |  |  |  |
| **Spine - BMD, g/cm<sup>2</sup> (mean (SD))** | 0.750 (0.155) | 0.735 (0.152) | 0.731 (0.117) | 0.771 (0.174) | 0.770 (0.176) | 0.673 |
| **Spine - Tscore (mean (SD))** | -2.125 (1.386) | -2.261 (1.361) | -2.304 (1.038) | -1.930 (1.546) | -1.957 (1.572) | 0.659 |
| **Total hip - BMD, g/cm<sup>2</sup> (mean (SD))** | 0.623 (0.122) | 0.654 (0.101) | 0.673 (0.100) | 0.594 (0.133) | 0.551 (0.128) | <0.001 |
| **Total hip - Tscore (mean (SD))** | -2.526 (1.221) | -2.206 (1.012) | -2.054 (1.026) | -2.804 (1.319) | -3.236 (1.270) | <0.001 |

### Longitudinal changes in bone turnover markers

TRACP 5b and P1NP showed marked early suppression after denosumab initiation, and this suppression was generally maintained during follow up across age groups. Similar patterns were observed in the treatment naive cohort (Supplementary Fig. S1a,b) and the overall cohort (Supplementary Fig. S1c,d), with no apparent age dependent difference. These descriptive findings indicate that denosumab induced suppression of bone turnover markers was broadly comparable across age groups.

### Association between baseline age and lumbar spine BMD response

In the treatment naive cohort, the multivariable model adjusted for sex, BMI, and eGFR showed that higher baseline age was significantly associated with lower lumbar spine %BMD gain after denosumab treatment (overall age by time interaction, p = 0.013; Fig. 2a). Model based estimates showed that older baseline age was associated with lower lumbar spine %BMD gain, with a 0.187 percentage point lower gain at 1 year and a 0.293 percentage point lower gain at 3 years for each additional year of age (1 year: β = −0.187, p = 0.006; 3 years: β = −0.293, p = 0.031; Supplementary Table 2). These findings suggest that older baseline age was associated with a modestly smaller lumbar spine BMD gain during denosumab treatment in the treatment naive cohort. In the overall cohort, baseline age was not significantly associated with lumbar spine %BMD gain at either 1 or 3 years (Supplementary Fig. S2a and Supplementary Table 2).

**Figure 2.**
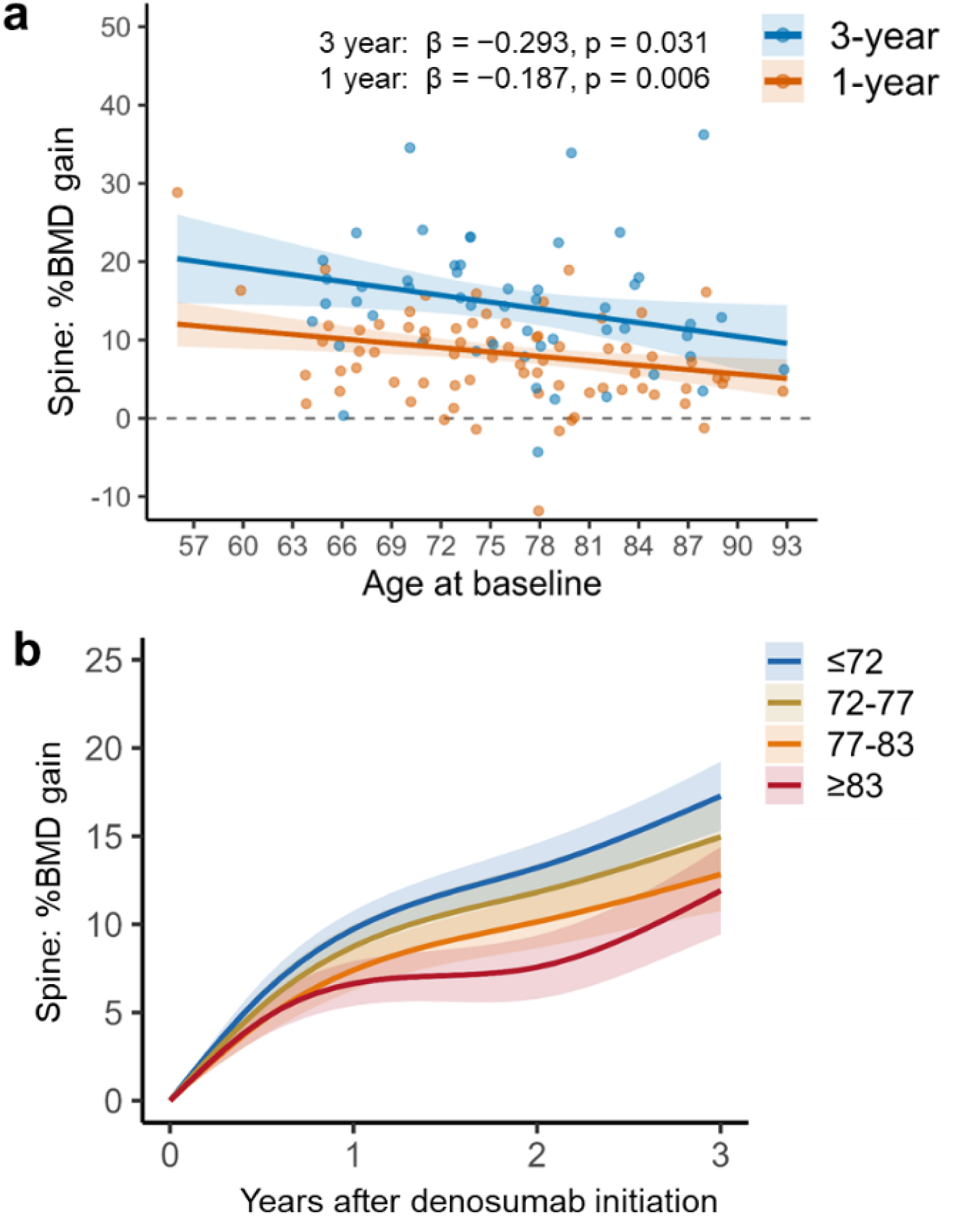
Association between baseline age and lumbar spine BMD response in the treatment naive cohort. a, Association between baseline age and lumbar spine %BMD gain at 1 and 3 years after denosumab initiation. Dots represent individual patients, and lines indicate model based estimates from multivariable linear mixed effects models. β values indicate the adjusted difference in %BMD gain per 1 year increase in baseline age. b, Model based trajectories of lumbar spine %BMD gain through 3 years after denosumab initiation by age group. Patients were stratified into four age groups: ≤72, 72 to 77, 77 to 83, and ≥83 years. Shaded areas indicate 95% confidence intervals.

To complement the continuous age analysis, patients were divided into four age groups based on cohort specific quartiles of baseline age. Model based trajectories showed that lumbar spine %BMD gain increased over time across all age groups in both the treatment naive cohort and the overall cohort. In the treatment naive cohort, the youngest age group showed numerically greater 3 year %BMD gain than the oldest age group (≤72 years: 17.8% versus ≥83 years: 12.6%; Fig. 2b). In the overall cohort, the age group trajectories were more closely aligned, with no clear age dependent separation (Supplementary Fig. S2b). These findings were consistent with the continuous age analysis and illustrated longitudinal response patterns across age groups.

### Association between baseline age and total hip BMD response

In the treatment naive cohort, baseline age was not significantly associated with total hip %BMD gain at 1 or 3 years after denosumab initiation (1 year: β = −0.011, p = 0.826; 3 years: β = 0.028, p = 0.727; Fig. 3a and Supplementary Table 3). Consistent with this, model-based age group trajectories showed longitudinal increases in total hip %BMD gain across age groups, without a clear pattern of attenuated response in older patients (Fig. 3b).

**Figure 3.**
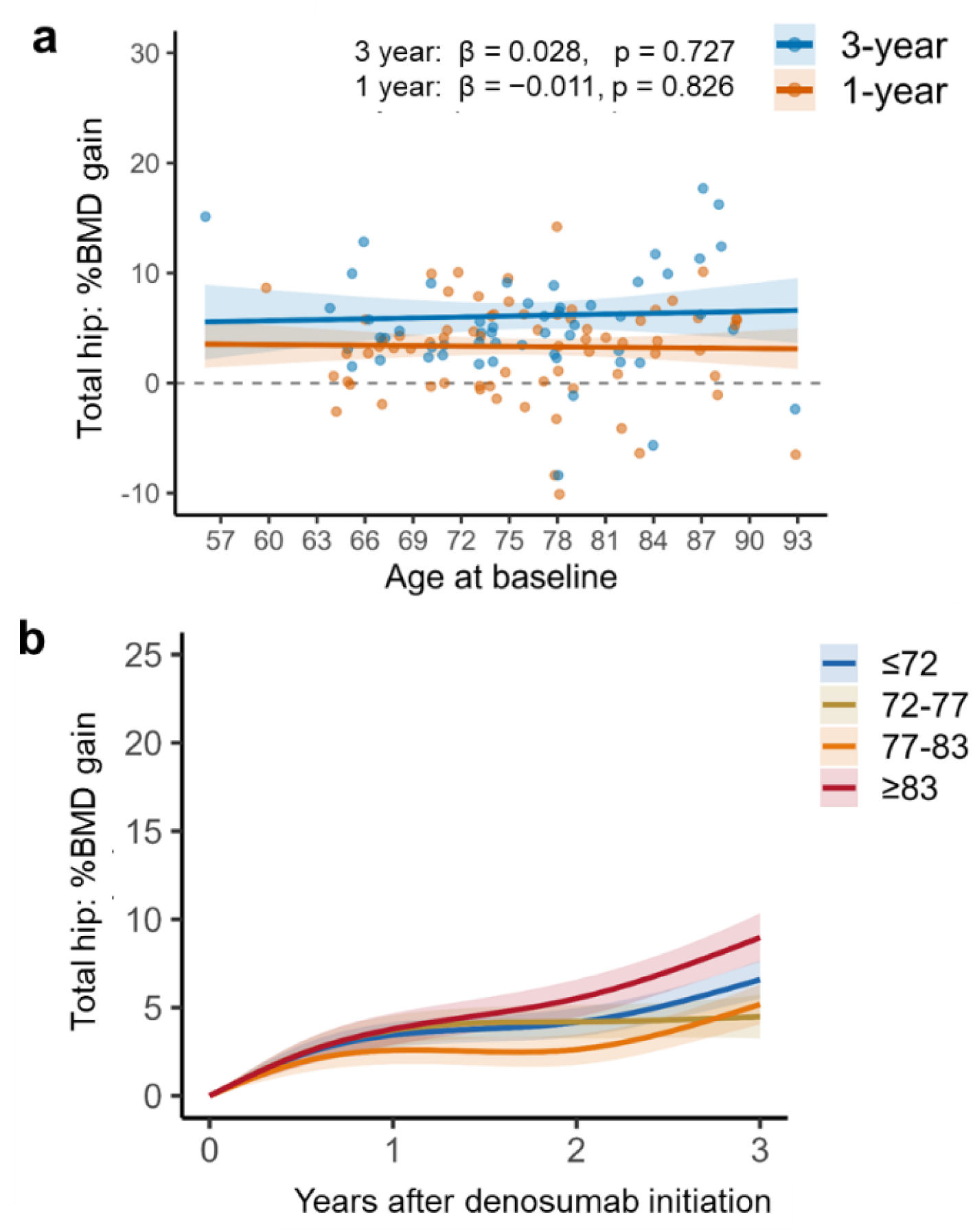
Association between baseline age and total hip BMD response in the treatment naive cohort. a, Association between baseline age and total hip %BMD gain at 1 and 3 years after denosumab initiation. Dots represent individual patients, and lines indicate model based estimates from multivariable linear mixed effects models. β values indicate the adjusted difference in %BMD gain per 1 year increase in baseline age. b, Model based trajectories of total hip %BMD gain through 3 years after denosumab initiation by age group. Patients were stratified into four age groups: ≤72, 72 to 77, 77 to 83, and ≥83 years. Shaded areas indicate 95% confidence intervals.

In the overall cohort, time point specific age trends at 1 and 3 years were not statistically significant (1 year: β = −0.008, p = 0.787; 3 years: β = 0.127, p = 0.052; Supplementary Fig. S3a and Supplementary Table 3). Age group trajectories in the overall cohort also showed no clear age dependent separation (Supplementary Fig. S3b). Overall, total hip BMD gain appeared broadly preserved across age, with no consistent evidence of an attenuated response in older patients during denosumab treatment.

## Discussion

The main finding of this study was that older patient age was associated with a modestly smaller lumbar spine BMD gain after denosumab treatment in treatment naive patients, whereas no significant age related association was observed at the total hip. This association was not evident in the overall cohort, suggesting that the influence of age on denosumab associated BMD gain is limited, but may be detectable at the lumbar spine in patients without prior osteoporosis treatment.

The observed association between older age and smaller lumbar spine BMD gain in treatment naive patients may reflect age related changes in skeletal remodeling and bone formation capacity^9,11,23^. Denosumab primarily acts by inhibiting RANKL mediated osteoclast activity, leading to reduced bone resorption and increased BMD. However, the magnitude of BMD gain after antiresorptive therapy may also depend on baseline remodeling activity and the capacity of the bone microenvironment to respond over time. Aging is associated with reduced osteoblast function, altered coupling between resorption and formation, and changes in marrow and skeletal microarchitecture^23–27^. These age-related changes could blunt the apparent gain in BMD at the lumbar spine, even when bone resorption is effectively suppressed. In our cohort, both TRACP-5b and P1NP decreased after denosumab initiation across age groups. Thus, the modestly smaller lumbar spine response in older patients is unlikely to be explained solely by inadequate suppression of bone turnover, as reflected by these markers.

The site-specific pattern of our findings is clinically informative. Older age was associated with a smaller lumbar spine BMD gain in treatment naive patients, whereas total hip BMD gain was broadly preserved across age groups. This preserved hip response may partly reflect the potent effects of denosumab on cortical bone, including suppression of intracortical remodeling and reduction of cortical porosity^28,29^. Unlike bisphosphonates, denosumab does not bind to bone and may more readily access intracortical remodeling spaces, which may contribute to its cortical effects^30^. In addition, the lower baseline total hip BMD observed in the oldest patients may have contributed to the results. Given the clinical importance of hip fracture prevention in older adults, the preserved hip BMD response observed in this study may underscore the clinical value of denosumab in this population.

An important observation was that the age-related association seen in the treatment naive cohort was not evident in the overall cohort. Prior osteoporosis treatment may have modified subsequent BMD response to denosumab and obscured age-related differences. Patients who previously received osteoporosis medications may start denosumab with different bone remodeling states and residual drug effects from prior therapy.

These factors can influence the magnitude and timing of BMD change after switching to denosumab. Therefore, the treatment naive cohort may provide a clearer assessment of the relationship between age and the intrinsic response to denosumab. By contrast, the overall cohort may better reflect real world clinical practice, where patients often receive denosumab after other osteoporosis medications. These findings suggest that denosumab remains effective across a wide age range, including in patients with different prior treatment histories.

This study has several strengths. We evaluated longitudinal BMD responses for up to 3 years, which provides relatively longer treatment among real-world studies. We also analyzed baseline age as a continuous variable using mixed effects models, while complementing this approach with age group-based trajectory analyses. In addition, we performed separate analyses in treatment naive patients and in the overall cohort, allowing us to distinguish a cleaner initial treatment response from a broader real-world population with prior medication exposure.

Several limitations should be acknowledged. First, this was a retrospective observational study. Second, the sample size was modest, particularly after stratification by age group and treatment history. Third, BMD was assessed using DXA, which does not capture bone microarchitecture or bone strength and may be affected by osteophytes and vascular calcification^31,32^. Finally, fracture outcomes were not evaluated as a primary endpoint due to limited sample size.

In conclusion, baseline age showed a modest, site-specific association with denosumab related BMD gain. In treatment naive patients, older age was associated with smaller lumbar spine BMD gain, whereas total hip BMD response was preserved across age. These findings suggest that denosumab provides BMD benefits across older age groups, while the magnitude of response may vary by skeletal site and treatment history. Together, these results may help clinicians incorporate patient age into denosumab treatment decisions in routine clinical practice.

## Data Availability

Aggregate data supporting the findings of this study are included in the manuscript and supplementary materials. Patient level data may be made available from the corresponding author upon reasonable request and with appropriate institutional approv

## Acknowledgments

We thank Ayano Oyamada, Kei Gonsho, Miho Mochizuki, and Tomoko Towatari for their cooperation in collecting clinical data.

## Author contributions

K.I. conceived and designed the study, collected and analyzed the data, interpreted the results, led the study, and wrote the initial draft. T.A. performed the statistical analysis and contributed to the initial draft. C.M., M.I., T.N., P.V., and S.T. contributed to data analysis and interpretation. W.R., P.P., and B.A.A. provided critical feedback and revised the manuscript. All authors reviewed and approved the final manuscript.

## Supplementary File

**Supplementary Figure S1.**
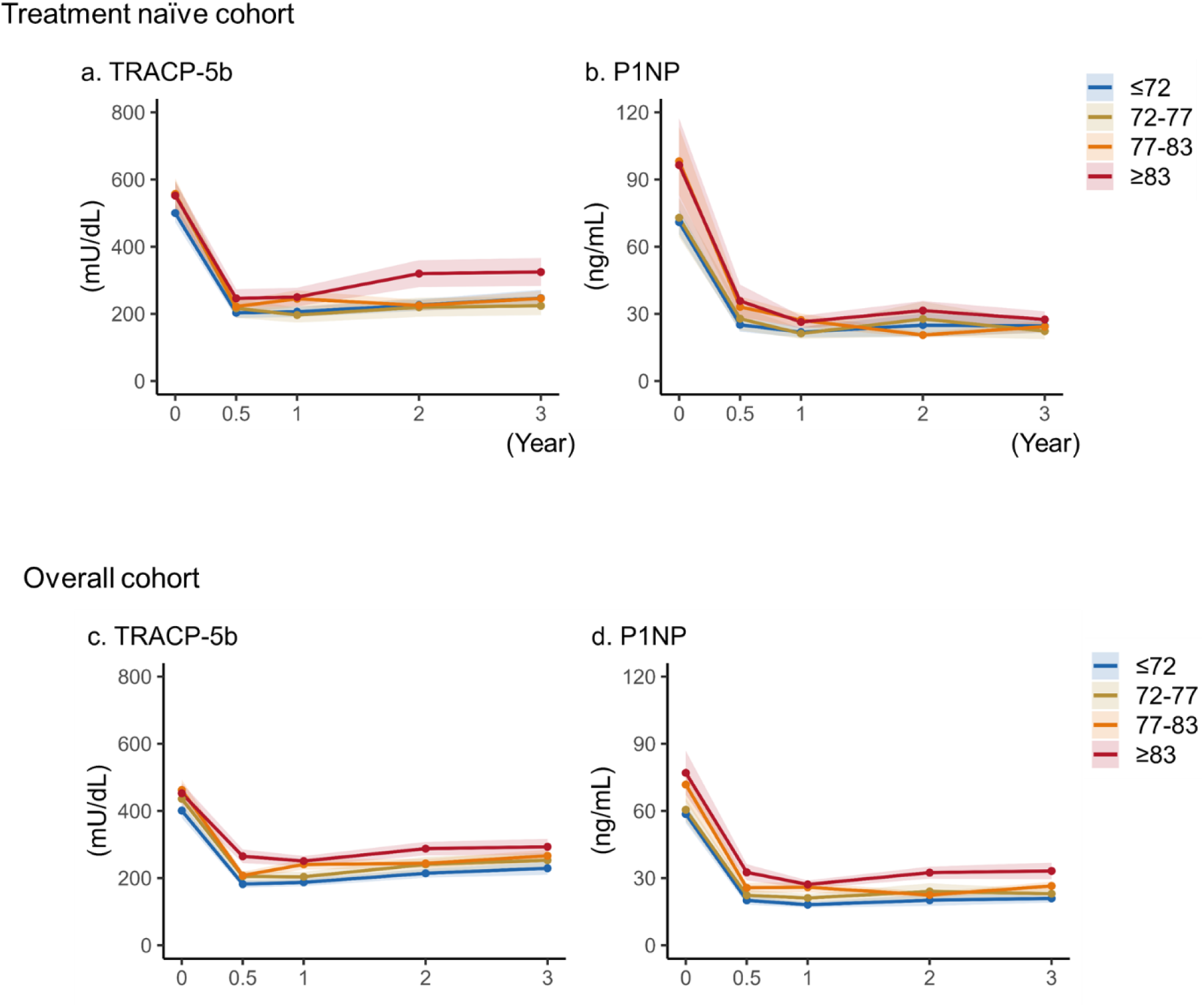
Changes in bone turnover markers after denosumab initiation by age group. a, Longitudinal changes in serum TRACP-5b levels in the treatment naive subset. b, Longitudinal changes in serum P1NP levels in the treatment naive subset. c, Longitudinal changes in serum TRACP-5b levels in the overall cohort. d, Longitudinal changes in serum P1NP levels in the overall cohort. Patients were stratified into four age groups: ≤72, 72 to 77, 77 to 83, and ≥83 years. Values are shown at baseline and 0.5, 1, 2, and 3 years after denosumab initiation. Lines and points indicate mean values, and shaded areas indicate SD.

**Supplementary Figure S2.**
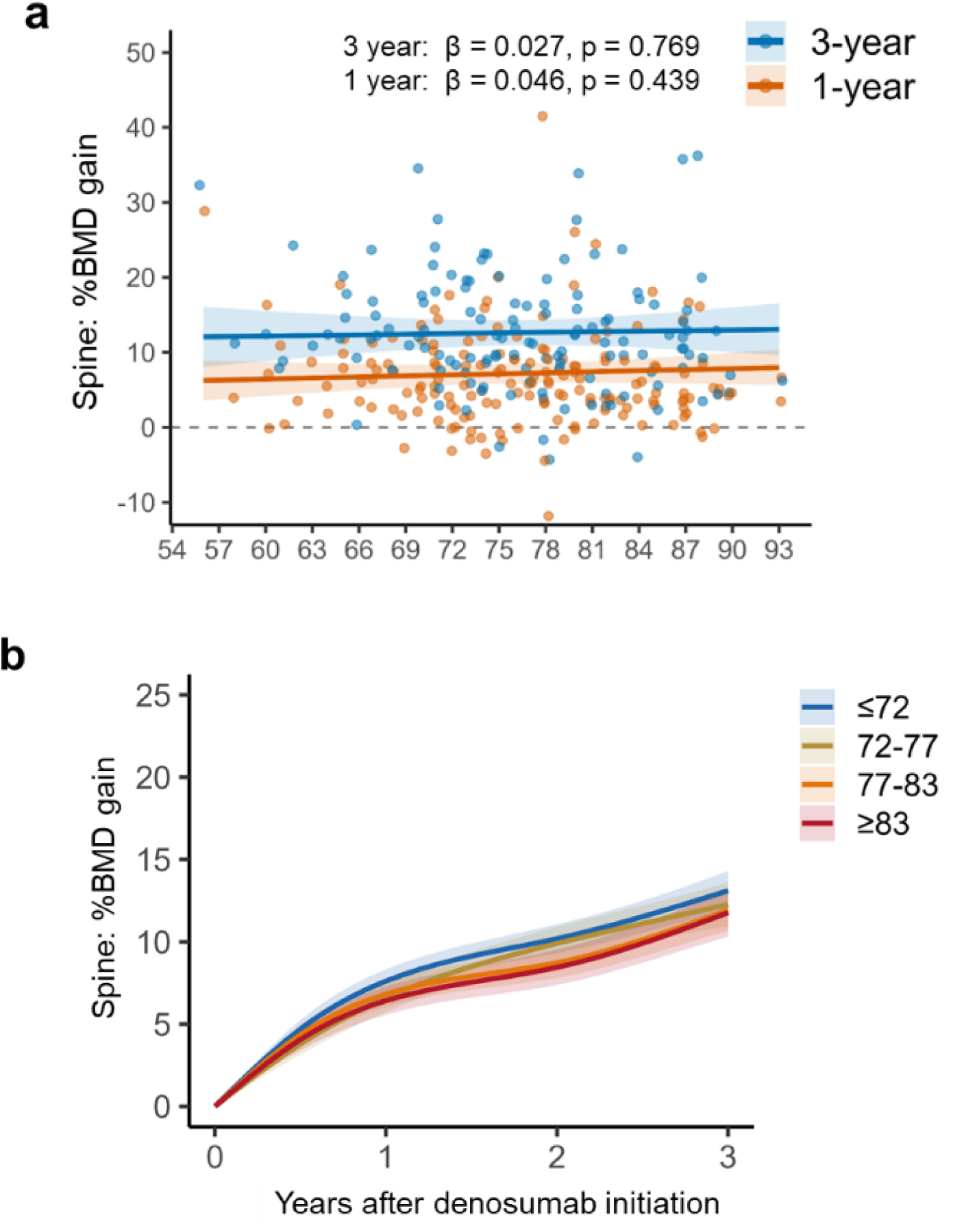
Association between baseline age and lumbar spine BMD response in the overall cohort. a, Association between baseline age and lumbar spine %BMD gain at 1 and 3 years after denosumab initiation in the overall cohort. Dots represent individual patients, and lines indicate model based estimates from multivariable linear mixed effects models. β values indicate the adjusted difference in %BMD gain per 1 year increase in baseline age. b, Model based trajectories of lumbar spine %BMD gain through 3 years after denosumab initiation by age group in the overall cohort. Patients were stratified into four age groups: ≤72, 72 to 77, 77 to 83, and ≥83 years. Shaded areas indicate 95% confidence intervals.

**Supplementary Figure S3.**
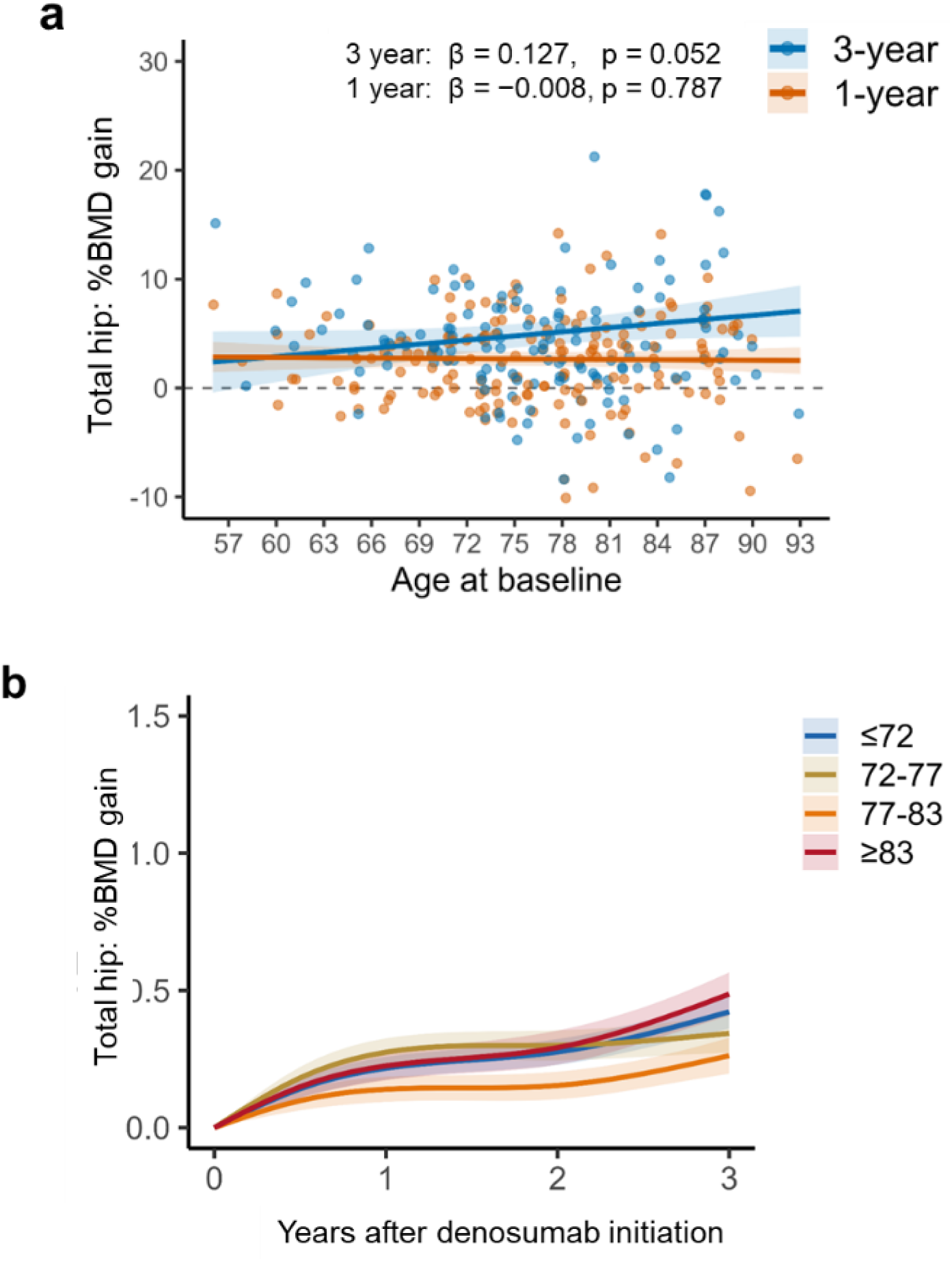
Association between baseline age and total hip BMD response in the overall cohort. a, Association between baseline age and total hip %BMD gain at 1 and 3 years after denosumab initiation in the overall cohort. Dots represent individual patients, and lines indicate model based estimates from multivariable linear mixed effects models. β values indicate the adjusted difference in %BMD gain per 1 year increase in baseline age. b, Model based trajectories of total hip %BMD gain through 3 years after denosumab initiation by age group in the overall cohort. Patients were stratified into four age groups: ≤72, 72 to 77, 77 to 83, and ≥83 years. Shaded areas indicate 95% confidence intervals.

**Supplementary Table 1.**
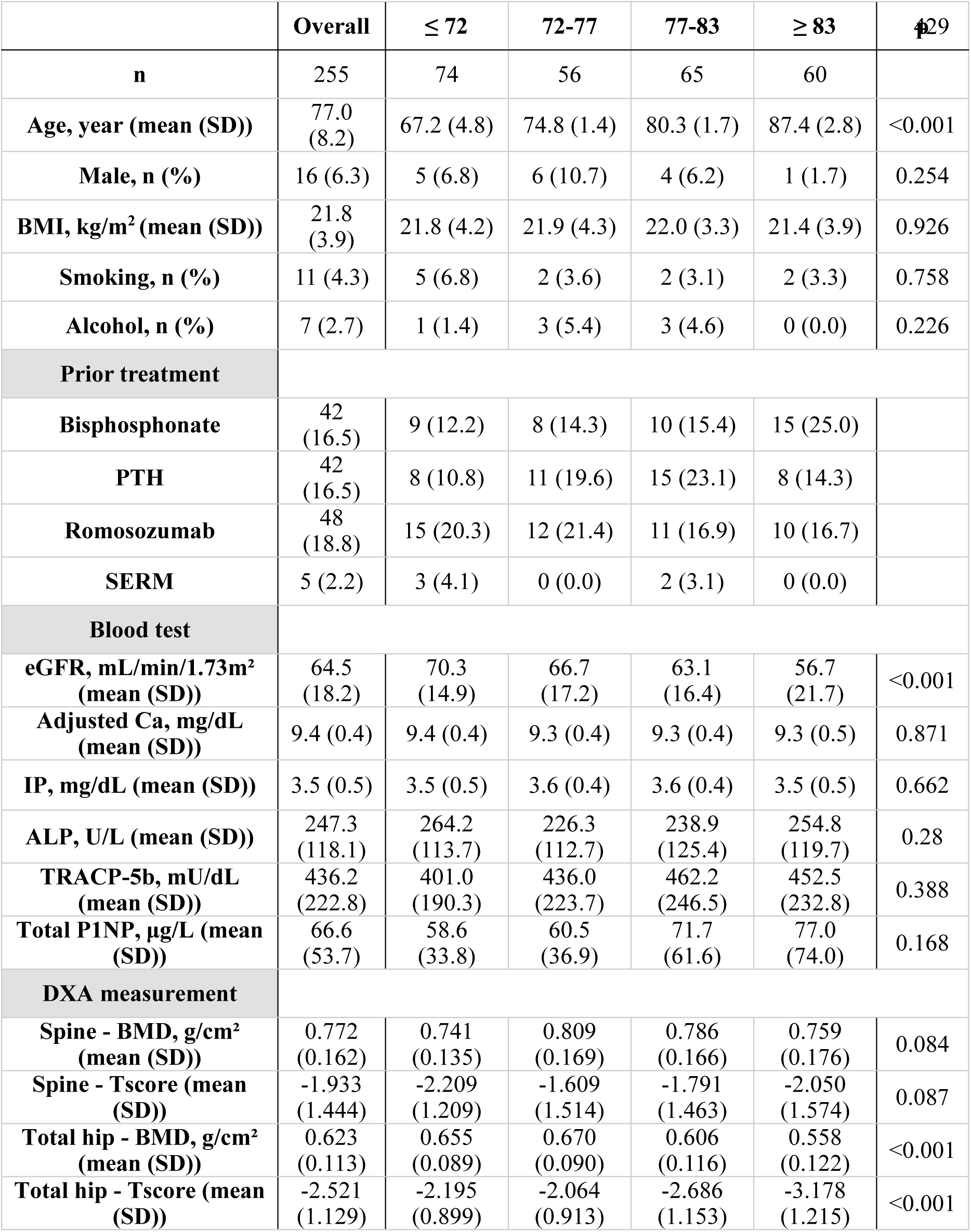
Demographic Data in overall cohort. PTH = parathyroid hormone analog; SERM = selective estrogen receptor modulator; TRACP-5b = Tartrate-resistant acid phosphatase-5b; P1NP = procollagen type 1 N-terminal propeptide; IP = inorganic phosphate; ALP = Alkaline phosphatase; Adjusted Ca=albumin-corrected calcium; DXA = Dual-energy X-ray absorptiometry; BMD = bone mineral density.

**Supplementary Table 2.** Multivariable model results for lumbar spine %BMD gain at 1 and 3 years. β values indicate the adjusted difference in lumbar spine %BMD gain per 1 year increase in baseline age. Models were adjusted for sex, BMI, and eGFR in the treatment naive cohort, and for sex, BMI, eGFR, and prior osteoporosis treatment status in the overall cohort. CI = confidence interval; BMD = bone mineral density; eGFR = estimated glomerular filtration rate.

|  | Treatment naïve cohort |  | Overall cohort |  |
| --- | --- | --- | --- | --- |
| | $\beta$ (95% CI) | P value | $\beta$ (95% CI) | P value |
| **Year 1** | -0.187 [-0.320, -0.056] | 0.006 | 0.046 [-0.071, 0.164] | 0.439 |
| **Year 3** | -0.293 [-0.559, -0.027] | 0.031 | 0.027 [-0.153, 0.208] | 0.767 |

**Supplementary Table 3.** Multivariable model results for total hip %BMD gain at 1 and 3 years. β values indicate the adjusted difference in total hip %BMD gain per 1 year increase in baseline age. Models were adjusted for sex, BMI, and eGFR in the treatment naive cohort, and for sex, BMI, eGFR, and prior osteoporosis treatment status in the overall cohort. CI = confidence interval; BMD = bone mineral density; eGFR = estimated glomerular filtration rate.

|  | Treatment naïve cohort |  | Overall cohort |  |
| --- | --- | --- | --- | --- |
| | $\beta$ (95% CI) | P value | $\beta$ (95% CI) | P value |
| **Year 1** | -0.011 [-0.113, 0.090] | 0.826 | -0.008 [-0.070, 0.053] | 0.787 |
| **Year 3** | 0.028 [-0.133, 0.190] | 0.727 | 0.127 [-0.001, 0.255] | 0.052 |

